# Effects of Ramadan Fasting on Children and Young Adults with Type 2 Diabetes during the COVID-19 Pandemic: A Prospective Study

**DOI:** 10.1101/2025.11.03.25339372

**Authors:** Hala K. Elmajnoun, Shaun Gorman, Suma Uday, James E Greening, Wojood Bensaid, Abu-Bakr Abu-Median, Parvez I. Haris

## Abstract

**Objectives:** The COVID-19 pandemic created unique circumstances for Ramadan fasting (RF) that have not been previously experienced in recent history. This study aimed to explore the impact of RF on lifestyle patterns, mental health, and glycaemic control among young people with type 2 diabetes (T2D) during the COVID-19 pandemic in the UK.

**Methods:** A prospective, observational, crossover, pilot study was conducted. The study participants included children and young adults with T2D aged 12-24 years, who practised RF for a minimum of 10 days. The study was conducted in three diabetes centres in the UK from March 2021 to June 2021. SoGoSurvey software was used to design online questionnaires that were used to collect data, including demographic information, medical history, impacts of the COVID-19 pandemic (weight and mental health) and lifestyle (diet patterns, physical activities and sleeping patterns). They were completed by the target age group 2 weeks before and 2 weeks after the month of Ramadan. Statistical analyses were performed using SPSS. Ethical approvals were obtained from the Health Research Authority (HRA) and from the Health and Life Science Faculty Research Ethics Committee at De Montfort University.

**Results:** Nine participants with T2D, including 7 females and 2 males aged from 14-22 years old (Mean ± SD; 17±3). Half of the participants (N=4) fasted safely the whole month of Ramadan. Participants reported that they fasted because of the health benefits (N=7) and they felt better during fasting (N=4). The glucose parameters, including HbA1c (P = 0.715), weight (P = 0.343), and body mass index (BMI) (P = 0.249), showed no statistically significant difference before and after RF. Most participants (N=8) were less active during fasting, and an altered sleeping pattern was reported (N=4). RF was associated with increased consumption of fruits and vegetables (N=4), decreased bread consumption (N=3) and an increase in consumption of desserts (N=3). In the period before RF, participants (N=4) reported that the COVID-19 pandemic was associated with weight gain, and this was not noticed during RF. Deterioration in mental health issues before RF (stress; N=3, emotional issue; N=3, depression; N=2) was reported by participants who had weight gain.

**Conclusion:** Young people, including children with T2D in the UK, fasted the whole month of Ramadan with no complications. Fasting during the COVID-19 pandemic was associated with a variable diet and improvement in mental health in study participants with T2D. Larger studies are necessary to determine the importance of education sessions before RF in the UK.

## Background

Fasting during the month of Ramadan is obligatory for all Muslims who have reached the age of puberty. During Ramadan fasting (RF), millions of Muslims worldwide abstain from food and drink from dawn to sunset every day for a continuous period of 29 to 30 days; once a year, in the 9th month of the lunar calendar [1, 2]. However, those who are ill or travelling are exempted from fasting. Despite these exemptions, many Muslims with chronic diseases, such as diabetes, fast during Ramadan. Over the last few decades, the impacts of RF on the biological modulation of plasma metabolites in patients with diabetes have been widely discussed. The findings, however, have been contradictory. Some researchers have noted that RF has a positive impact on weight loss, glycaemic control, and lipid profile in both healthy individuals and those with T2D [1]. In contrast, other studies indicate that RF has no significant effect on these metabolites [2]. The vast majority of the work in this area has focused on examining the effect of RF on blood glucose biomarkers among adults with T2D and healthy subjects [1, 2]. To date, no study has looked specifically at the effects of RF on children and young adults with T2D [3]. The incidence of T2D in children and young adults is continually rising all over the world, and about 42,000 newly diagnosed cases were reported in 2021 [4]. According to Diabetes UK, “there are more than 7,000 children and young adults under 25 with T2D in England and Wales” (Diabetes.co.uk, 2019). Moreover, in Manitoba, Canada, the incidence of T2D among children reached up to 20.55/100,000 [5]. This dramatic rise is a growing public health concern worldwide; thus, for the first time, the present study will take the opportunity to evaluate the effect of RF on the blood glucose biomarkers and diet patterns within these groups in the UK.

In addition to the unprecedented circumstances of the COVID-19 pandemic, the number of cases of diabetes and prediabetes is anticipated to double worldwide. Consequently, this may increase the incidence of morbidity and mortality among this group of people [6]. On the other hand, a systematic review conducted worldwide during May 2020 reported that children and young adults (up to 21 years old) with COVID-19 had a very good prognosis, and most of the cases recovered completely, even for people with pre-existing medical problems [7]. In terms of the impacts of the COVID-19 pandemic on children, it was reported that the mortality rate was about 0.09% among a total of around 8,000 confirmed cases [7]. This result was based on analysing data from healthy children and children with comorbidities [7]. Moreover, it has been noticed that most children confirmed with COVID-19 were asymptomatic [8]. However, severe to moderate symptoms have been recorded among infants and some children with chronic diseases [9].

According to Diabetes UK, children with diabetes can get COVID-19 infections; however, the risk of developing severe illnesses is extremely rare (Updates: Coronavirus and Diabetes, 2020). Nevertheless, these people are still vulnerable to COVID-19 infection, and careful precautions with close health care observations are highly recommended, particularly, patients with uncontrolled blood glucose and who have secondary complications of diabetes. Therefore, it is important to provide evidence on how these patients have been affected during the pandemic. Throughout the period of the COVID-19 pandemic, individuals with type 2 diabetes who were willing to fast were expected to take extra care to ensure that they were physically active and had sufficient sleep to better manage their diabetes [10]. The inability to carry out physical activity due to COVID-19 quarantine has been identified as a serious health issue, and some have suggested types of physical activity that people can perform at home during that time [11]. Thus, this study was conducted to provide an overview of how young people with T2D are being affected by the pandemic and whether they were physically active or not during the month of Ramadan.

There are several reports regarding the high prevalence of mental health problems linked to the COVID-19 pandemic, and some of these were attributed to the impacts on daily life [12, 13]. It has also been reported that the mental health of children and older people has been affected by the unprecedented circumstances of the pandemic [14, 15]. In addition, several studies in the literature reported improvements in mental health associated with RF [16, 17]. One study reported improvements in people with diabetes who experienced depression [18]. A systematic review reported that RF does not cause new mental health problems, and there were no major reasons to advise people not to fast, except for people with major psychiatric disorders [19].

Moreover, it has been shown that fasting has a favourable effect by helping to overcome stress and having a positive impact on alertness [20, 21]. Therefore, these impacts may have helped some Muslims who had experienced mild stress during the COVID-19 outbreak, such as anxiety due to social isolation and recurrent lockdown measures. Considering the current literature, this study hypothesised that fasting during Ramadan may bring positive health benefits for healthy people, but those with pre-existing health issues might develop serious complications by engaging in fasting without seeking medical consultation. Some people may benefit, while others may suffer.

### Research aims/objectives

#### Research aim

- Examine the impact of RF on lifestyle aspects, mental health, and blood glucose parameters in children and young adults (12 to 24 years old) with T2D during the COVID-19 pandemic in the UK.

#### Research objectives

- To investigate the impacts of RF on the glycosylated haemoglobin (HbA1c), body mass index (BMI), blood pressure (BP), and body weight (BW) in children and young adults with T2D (aged 12–24 years old), during the COVID-19 pandemic. A prospective online survey questionnaire will be designed to gather and compare these data before and after Ramadan 2021 in the UK.
- To examine the impacts of RF on diet patterns (food frequencies), physical activities, sleep patterns, and mental health issues among children and young adults with T2D during the pandemic. This will be evaluated by comparing data before and after Ramadan 2021 in the UK.
- To evaluate the impact of RF during the COVID-19 pandemic on the feasibility of accessing medical services and maintaining blood glucose control for children and young adults with T2D in the UK.

## Methods

### Study design and setting

This study was designed as a prospective, observational, crossover pilot study, meaning that the cases act as controls of themselves. The data collected before RF were compared with the data collected after RF. An online questionnaire survey was used to collect data for the study at two measurement points: two weeks before Ramadan and two weeks after Ramadan. The study was undertaken at three medical centres in the UK, including the Leicester Royal Infirmary (University Hospitals of Leicester NHS Trust), the Birmingham Children’s Hospital (Birmingham Women’s and Children’s NHS Foundation Trust), and the Bradford Royal Infirmary (Bradford Teaching Hospitals NHS Foundation Trust). The average fasting hours during Ramadan 2021 in the UK were between 16-18 hours per day, and the average temperature was around 15°C.

### Designing the questionnaire

The questionnaire was adapted from the EPIC-Norfolk food frequency questionnaire (FFQ) [22]. This Epic–Norfolk FFQ comes along with an Open Government Licence or public sector information, which will allow the copying, use, publishing, and transmission of information [22]. It was modified and converted to an online version. In addition, many questions related to the COVID-19 pandemic, patients with diabetes, physical activities, and sleeping patterns were added. This adaptation was carried out using SOGOSurvey software. The questions were carefully written and designed using clear, understandable lay language to increase the response rate and collect the precise information required to achieve the study’s aims. To avoid bias in responses, closed-ended questions such as yes/no, and multiple-choice questions, were used. The questionnaire was structured in a specific way to maintain the consistency and order of the required information. Starting with a small introduction to the study, the participants were guided to choose the suitable participant information sheets (PIS) according to their age. All the PISs were provided as links at the beginning of the questionnaire, where the respondents could easily access and read them at a convenient time. Four different versions of the PISs were developed for: 12–15-year-olds, parents, 16–18-year-olds, and 18+ (Supplementary Material). Subsequently, the interested participants were required to agree to all the statements on the consent form. Two types of consent forms were provided in the surveys, one for parents (for children from 12 to 15) and one for young adults (from 16 to 24) (Supplementary Material). The questionnaire was then divided into four sections:

1. Information about you: age, gender, employment status (for people aged 16 to 24 years old), and physical measurements.
2. Ramadan fasting and COVID-19: in this section, the participants must answer some questions related to COVID-19 and fasting.
3. Health and medical history: this section provided background on the study participant’s medical history (in relation to diabetes) and the level of blood glucose control before and after the month of Ramadan.
4. 4. Lifestyle-related information: this section covers all questions related to diet intake, sleep duration/time, and physical activity. Also, some questions related to COVID-19 were added to explore the impact of COVID-19 on all aspects of lifestyle.

The participants were asked to do all these steps before and after the month of Ramadan by two weeks. SOGOSurvey was used to create several survey URL links and share them with interested participants before and after the month of Ramadan. The contact with the participants was indirect, either by email or over the phone. The links to the questionnaires were sent by email and text message according to the participant’s request. The participants were provided with two links to access; children aged 12 to 15 years old received two links, one before the month of Ramadan and one after Ramadan. The two other links were sent to study participants aged 16 to 24 years old: one before and one after Ramadan. PDF copies of all the survey links are provided in the Supplementary Material. No personal information was collected in the questionnaire. The survey was self-administered, and the participants were instructed that they could complete all the survey sections within 15 to 20 minutes. However, the researcher helped some participants complete the survey when they had some technical problems due to the survey links. Children aged 12 were informed that they can complete it with their parents in order to help them answer all the questions as precisely as possible.

To ensure the validity and reliability of the designed questions, the questionnaire was distributed among different age groups, and they were asked to provide evaluation/feedback, such as the time taken to complete the survey and the clarity of the questions. The face validity technique was used [22]. All the feedbacks were taken into consideration to enhance the questionnaire further and avoid any confusion that might occur.

### Inclusion and Exclusion criteria

This study included participants from <u>a</u>ll ethnic groups in the UK who were able to read and understand basic/simple English language. The target age group was children and young adults, from 12 to 24 years old. Moreover, participants who were diagnosed with T2D and decided to fast for a minimum of 10 days during the month of Ramadan were included. However, people aged less than 12 years old and more than 24 years old were excluded. In addition, this study did not include people with cognitive impairment, individuals who do not know/understand the English language, and participants who decided to withdraw from the study after completing only the questionnaire before the month of Ramadan.

### Sample size and sampling method

The sample size was calculated using G*Power for the two means of the same group, and it is assumed that a two-tailed test statistical analysis was used. The power was 80%, which is the minimum power in medical research studies [23]. The effect size (the significant difference between two means before and after the month of Ramadan) and the power (the ability to detect differences between two means) were selected based on the study design. This study involved two sampling methods: convenience sampling (Non-Probability Sampling Methods) and cluster sampling (Probability Sampling Methods). Convenience sampling was used as the easiest and simplest way to get more participants. Also, cluster sampling was used to divide the individuals into subgroups according to their age group and the NHS location, as they could be identified as clusters. The sample size was adjusted based on alternative methods, such as previous similar or analogous studies in the literature [23]. The average sample size of pilot studies among RF varies from 8 to 52 participants among healthy adults and patients with diabetes [24, 25].

### Gaining the REC and HRA approvals

This research study complied with the Medical Research Council’s (MRC) research ethics guide and the National Ethics Guidelines for Biomedical Research Involving Children. Ethical approvals were obtained locally from the Research Ethics Committee (REC), Health Research Authority (HRA) in England, UK, and Health and Life Science Faculty Research Ethics Committee at De Montfort University, Leicester, UK.

### Recruitment and data collection methods

The recruitment started one month before the start of Ramadan 2021. On each site, the interested participants were first approached by the gatekeepers, who invited them to participate in the study. They have been contacted by phone and/or email, and they received the research flyers and the PISs to have more information about the study. Then, the participants who were interested in the research were guided to contact the researcher, who provided them with all the information needed and guided them throughout the course of the study. To increase the response rate, the interested participants were reminded a few times to complete the questionnaires.

## Data analysis methods

Data were exported from the SOGOSurvey software to the Statistical Package for the Social Sciences (SPSS), version 26 (IBM Corporation, Armonk, NY, USA), for analysis. In SPSS, the data were organised, cleaned, and coded. Both anonymisation and pseudonymisation techniques were used during data analysis. The survey questions were divided into three types: categorical variables, ordinal variables, and continuous variables. The distribution of continuous variables was examined using visual inspection and statistical tests, including Shapiro & Wilk’s test (p< 0.05) and histogram, Q-Q plot, and Box plot. Percentages and counts were used for categorical variables. Descriptive statistical analysis was conducted; mean ± standard deviation (SD) was used for continuous variables, which are normally distributed, and the median and interquartile range (IQR) or range were used for the skewed data. Paired t-test and Wilcoxon Signed Ranks test were used to compare the differences between outcome variables before and after the month of Ramadan for paired parametric and non-parametric data, respectively. Statistical significance was set at P < 0.05 (two-tailed).

## Results

### Participants characteristics

Out of 26 recruited participants with T2D, only 9 (7 male, 2 female) completed all the study stages (22.5% response rate). The mean age group was at (mean & SD; 17±3), and about two-thirds of the respondents were obese and about one-fifth were overweight (**Table 1**). Only one respondent was an employee and did not work during the pandemic period. The other respondents were unemployed. Participants were diagnosed with T2D for a mean duration of 3±2 years. All the participants have a family history of T2D, and most of them were on oral medications (N=8), and half of these participants were on diet/exercise as part of their diabetes management. Out of 8 respondents, half of them (N=4) fasted the whole month of Ramadan 2021, as illustrated in **Figure 1**. Moreover, 7 participants had observed the previous months of Ramadan over the last three years.

**Figure 1:**
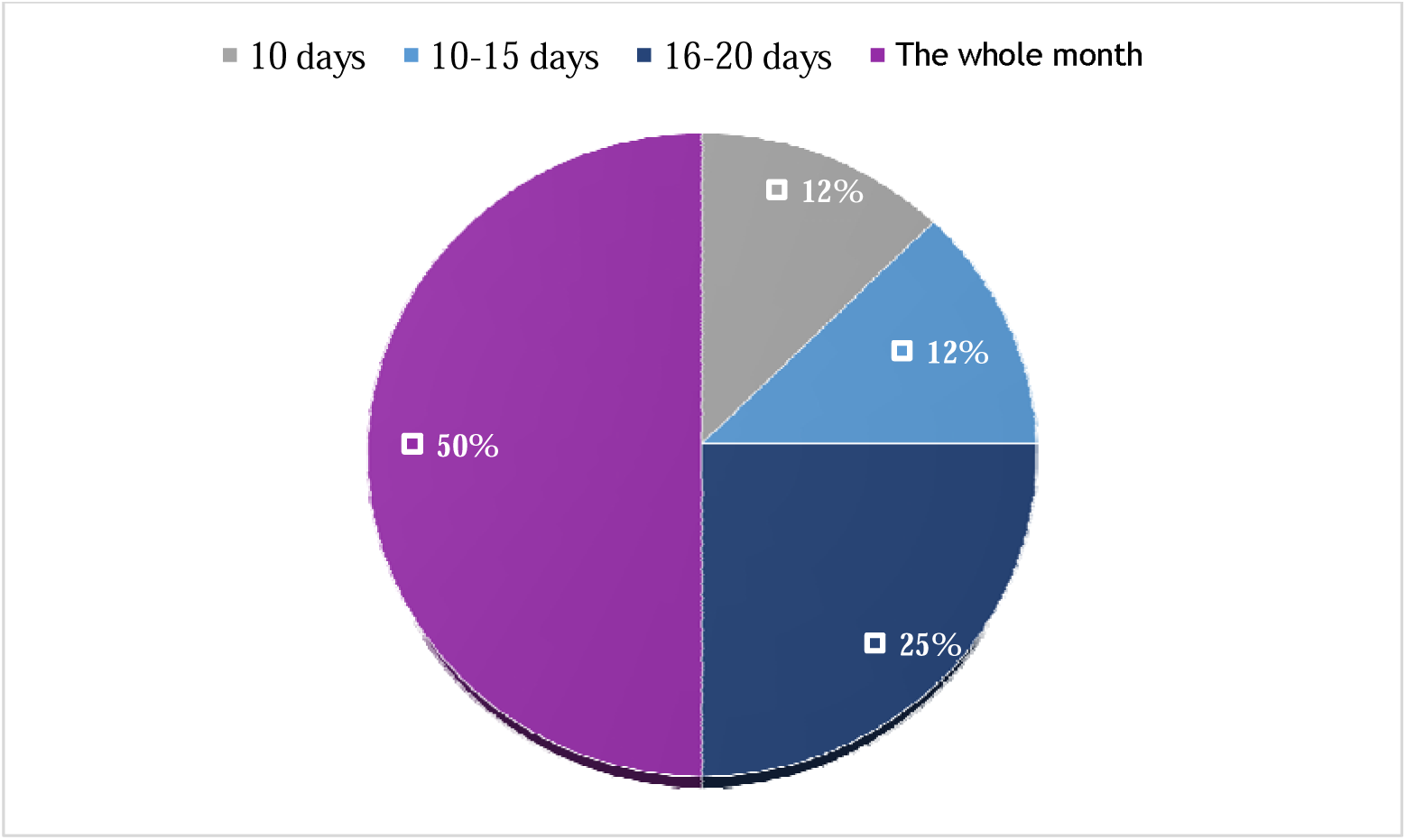
Fasting days during Ramadan 2021 (N=8). 50% (N=4); 25% (N=2); 12% (N=1). The whole month (30 days).

**Table 1:** Participant demographic characteristics, medical history and fasting during Ramadan (N=9)

| Participants characteristics | N |
| --- | --- |
| **Age; Mean ± SD; range** | 17±3 (14-22) |
| **Weight (Kg); Mean ± SD** | 87.67±19 |
| **Body mass index; Mean ± SD** | 30.81±7.47 |
| **Gender** |  |
| Male | 2 |
| Female | 7 |
| **BMI classification** |  |
| Healthy | 1 |
| Overweight | 2 |
| Obese | 6 |
| **Type 2 diabetes duration; Mean ± SD** | 3±2 |
| **Family history of T2D** |  |
| No | 0 |
| Yes | 9 |
| **Diabetes Medication** |  |
| Insulin injection | 0 |
| Oral medication | 8 |
| Insulin oral medication | 1 |
| With diet and exercise | 4 |
| **Fasting days (N=8)** |  |
| 10 days | 1 |
| 10-15 days | 1 |
| 16-20 days | 2 |
| The whole month | 4 |
| **Observing the previous months of Ramadan** |  |
| 2018 | 7 |
| 2019 | 8 |
| 2020 | 9 |
| **Employment status** |  |
| Employed | 1 |
| Unemployed | 4 |
| Not applicable | 4 |
| **working before Ramadan 2021 (during the COVID-19 pandemic)** |  |
| No | 4 |
| Not applicable | 5 |

**Table 2** shows that the vast majority of participants (N=7) did not have diabetes comorbidities, and 6 participants received diabetes education sessions during their illness. However, some participants reported that they have proteinuria (N=1) and pelvic inflammatory disease (N=1) as diabetes-associated comorbidities. Moreover, hypoglycaemia was reported by one study participant before the month of Ramadan. All participants reported that medical services were available before Ramadan 2021, and more than half (N=5) were in regular contact with their doctors. In addition, 5 participants reported that their diabetes was under control before Ramadan, and 7 participants reported that they were taking their medication regularly and that it had not changed for the month of Ramadan. Just two respondents reported that their medications were reversed (the usual morning dose was taken in the evening, and the usual evening dose was taken in the early morning) during the month of Ramadan. In addition, as shown in **Figure 2**, 7 respondents with T2D reported that they fasted during Ramadan because of the health benefits, and feeling better during fasting was reported by about half of these people (N=4). However, 2 respondents, aged 14 years and 15 years, stated that they fasted during the month of Ramadan due to social pressure.Dietary patterns among study participants with T2D during Ramadan 2021

**Figure 2:**
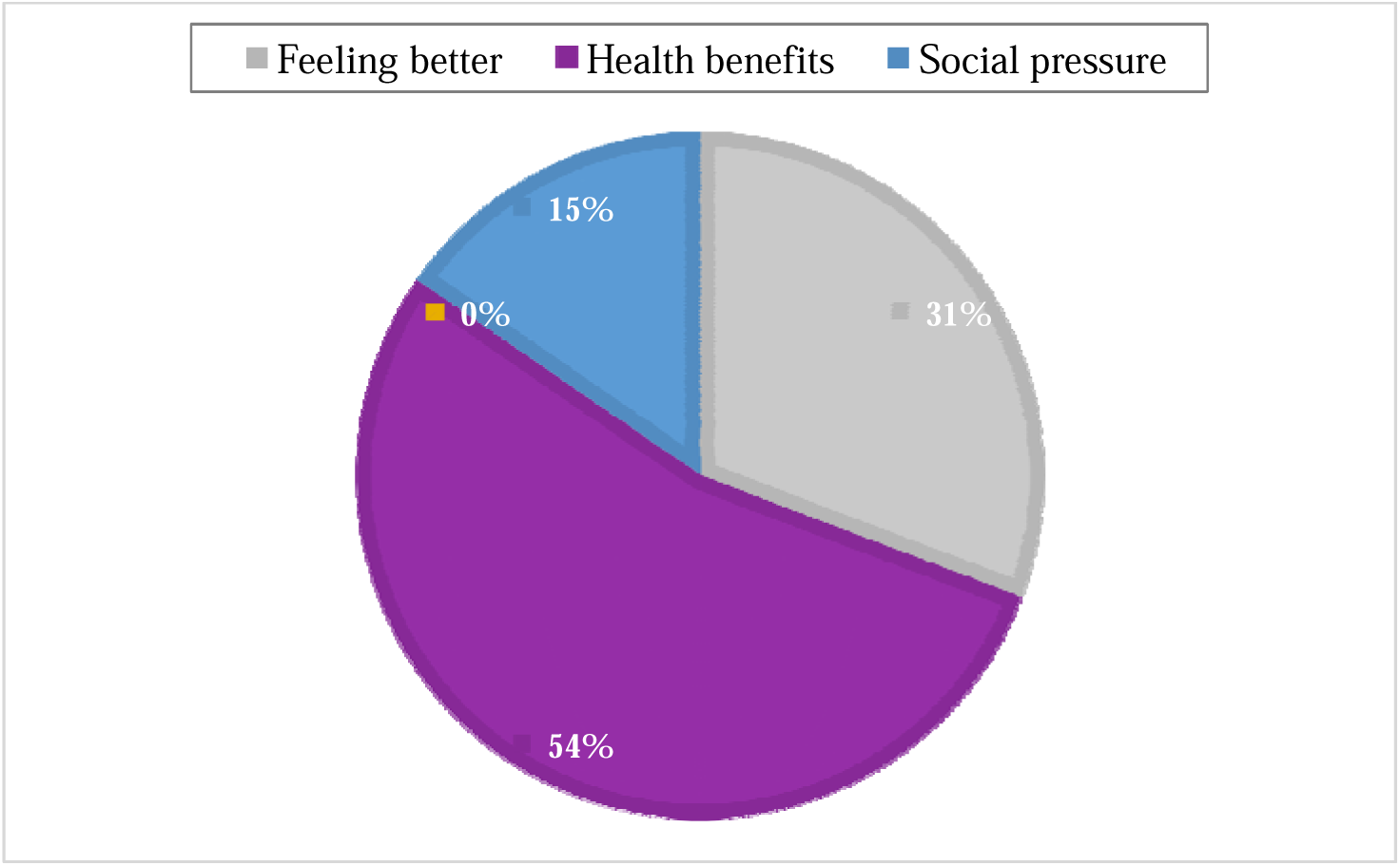
Reasons that motivated study participants to fast during the month of Ramadan (N=9). Percentages of responses: 54% (N=7); 31% (N=4); 15% (N=2).

**Table 2:** Medical characteristics of the respondents (N=9).

|  | N |
| --- | --- |
| **Diabetes associated comorbidities** |  |
| Heart disease | 0 |
| Kidney disease | 0 |
| Nerve damage | 0 |
| Vision | 0 |
| I do not know | 7 |
| High protein in urine | 1 |
| Pelvic inflammatory disease | 1 |
| **Diabetes symptoms before Ramadan 2021** |  |
| Hypoglycaemia | 1 |
| Hyperglycaemia | 0 |
| DKA | 0 |
| other | 0 |
| **Do you take your treatments regularly before Ramadan 2021?** |  |
| No | 2 |
| Yes | 7 |
| **Diabetes education** |  |
| No | 3 |
| Yes | 6 |
| **Change medications before Ramadan** |  |
| Reduced | 0 |
| Reversed (morning and evening medication) | 2 |
| Increased | 0 |
| None | 7 |
| **Diabetes control before Ramadan 2021** |  |
| It was under control | 5 |
| It was not under control | 0 |
| I cannot remember | 3 |
| **Medical services availability before Ramadan 2021** |  |
| No | 0 |
| Yes | 9 |
| **Regular contact with the doctor before Ramadan 2021** |  |
| No | 4 |
| Yes | 5 |
| **Reasons that motivate you to fast in Ramadan** |  |
| Health benefit | 7 |
| Feeling better | 4 |
| Social pressure | 2 |

**Table 3** shows the differences in dietary patterns before and during the month of Ramadan. The participants were asked to describe whether their diet was healthy, average (moderately healthy), or unhealthy. Out of 9 respondents, about two-thirds (N=6) reported that their regularly consumed diet was average before the month of Ramadan. The other third (N=3) reported that their diet was unhealthy, and this was similarly seen during the month of Ramadan. However, 2 respondents reported that they consumed healthy food during Ramadan (**Figure 3).** Most of the respondents reported that they consumed two meals per day before Ramadan and during Ramadan, at N=6 and N=5, respectively. However, one respondent had three meals during Ramadan **(Figure 4**). It was observed that participants consumed more fruits and vegetables during the month of Ramadan, from 2-3 portions/day (N=4) and 4 to 5 portions/day (N=2), compared to just one portion/day before the month of Ramadan (N=4). In addition, descriptive data indicated that there was a reduction in consumption of sugary drinks during the month of Ramadan for one respondent, from 6+ times per day to once a day. However, the consumption of desserts (cakes, doughnuts, and sweet biscuits) increased slightly from 1-3 times/month to 2-4 times/week in one third of the respondents (N=3) (**Table 3**).

**Figure 3:**
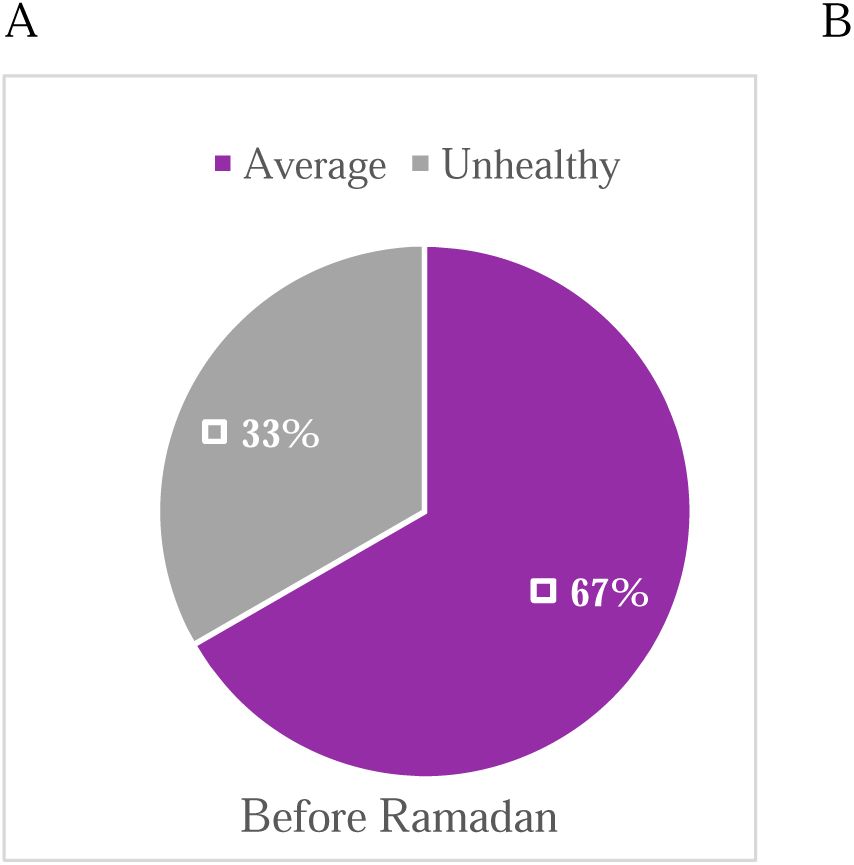
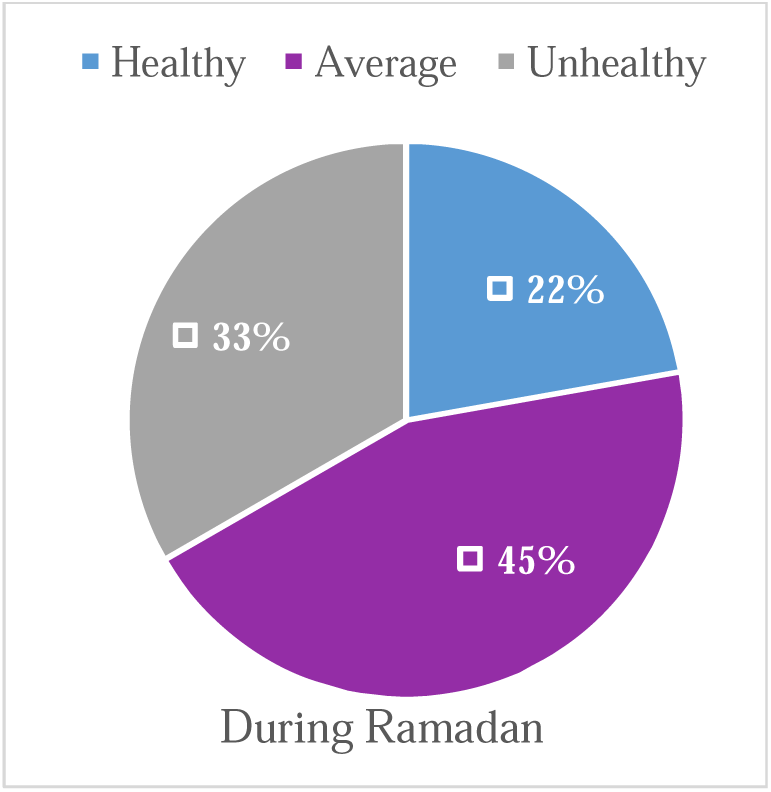
Comparing diet assessment for study participants with T2D between the regularly consumed diet before Ramadan (A) and the diet during the month of Ramadan (B) (N=9). 67% (N=6); 33% (N=3); 45% (N=4); 22% (N=2).

**Figure 4:**
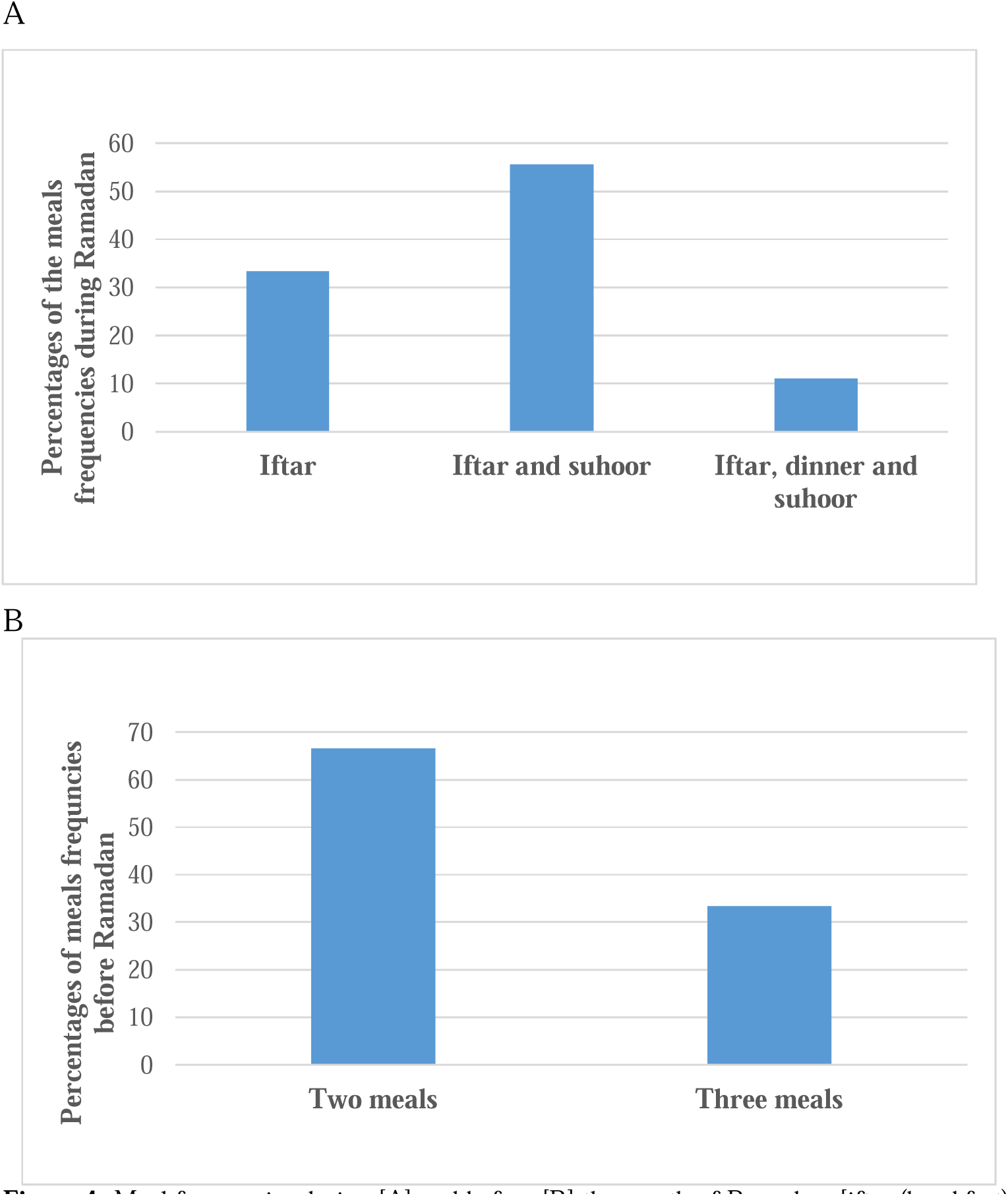
Meal frequencies during [A] and before [B] the month of Ramadan. [iftar (breakfast); suhoor (meal before dawn)], (N=9). 67% (N=6); 33% (N=3); 56% (N=5); 11% (N=1).

**Table 3:**
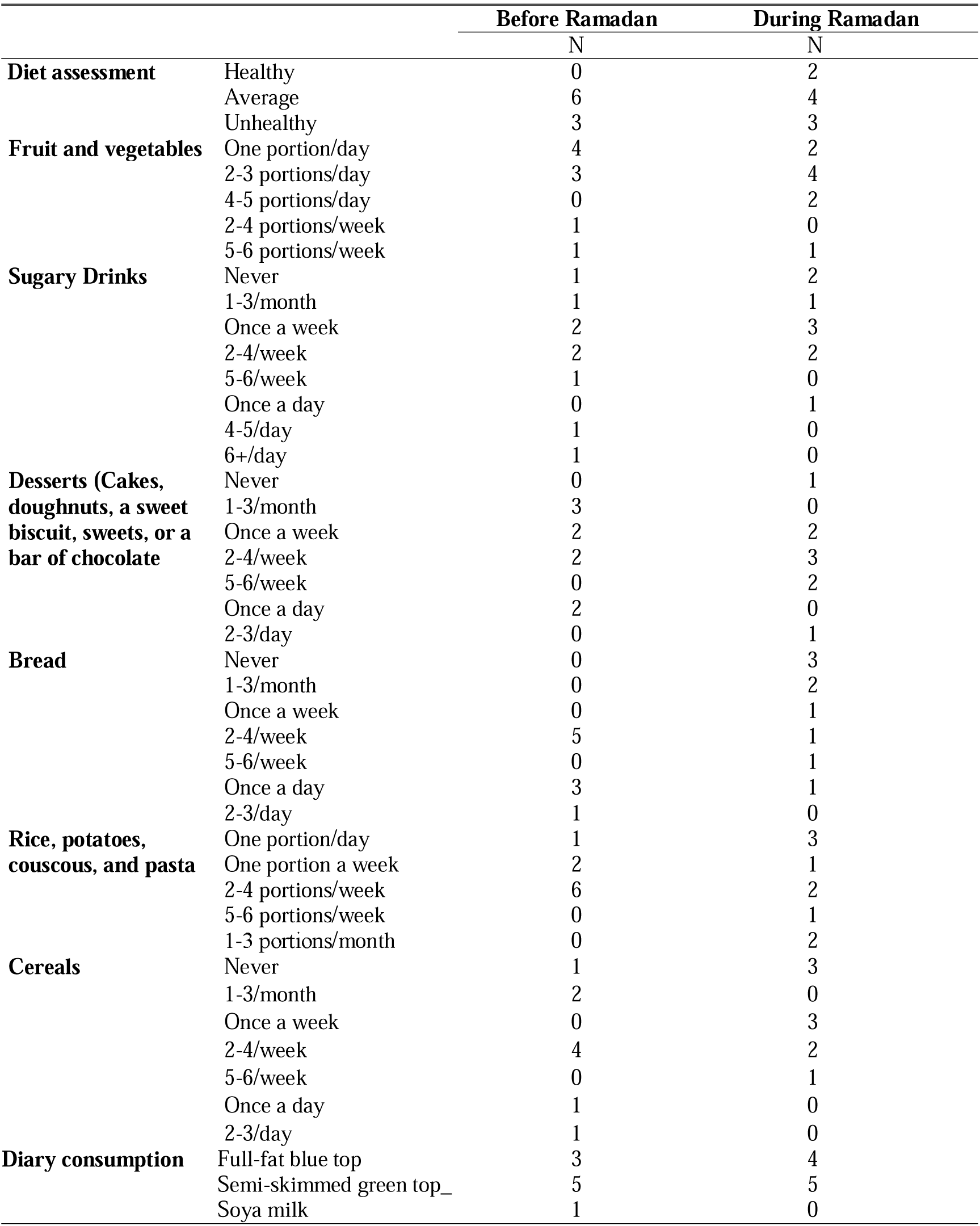
Comparison between the frequencies of the regularly consumed food before Ramadan and during the m onth of Ramadan (N=9).

Furthermore, 5 respondents reported that they usually consume bread 2-4 times per week before Ramadan. This was reduced to never (N=3) and 1-3/month (N=2) during the month of Ramadan (**Table 3**). This pattern was similarly observed for the consumption frequencies of cereals, where the consumption decreased from 2-4 times a week (N=4) before Ramadan to never (N=3) and once a week (N=3) during Ramadan. However, 6 participants reported that they usually eat starchy foods such as rice or potatoes in 2-4 portions per week before Ramadan, and 3 respondents reported that they consume one portion per day during Ramadan. However, the frequency pattern among the rest of the respondents varied from one portion a week to 1-3 portions a month (**Table 3**). Moreover, there were no differences in the usual dairy consumption before Ramadan and the frequency consumed during the month of Ramadan.

### Physical activities and sleeping patterns in T2D study participants during the month of Ramadan

Out of 9 respondents, 7 (78%) reported that they were practising exercise regularly before Ramadan, while only 4 (44%) practised regular exercise during Ramadan (**Figure 5**). The most common types of exercise were lifting weights (N=3) before Ramadan and brisk walking (N=4) during Ramadan, as illustrated in **Figure 6**.

**Figure 5:**
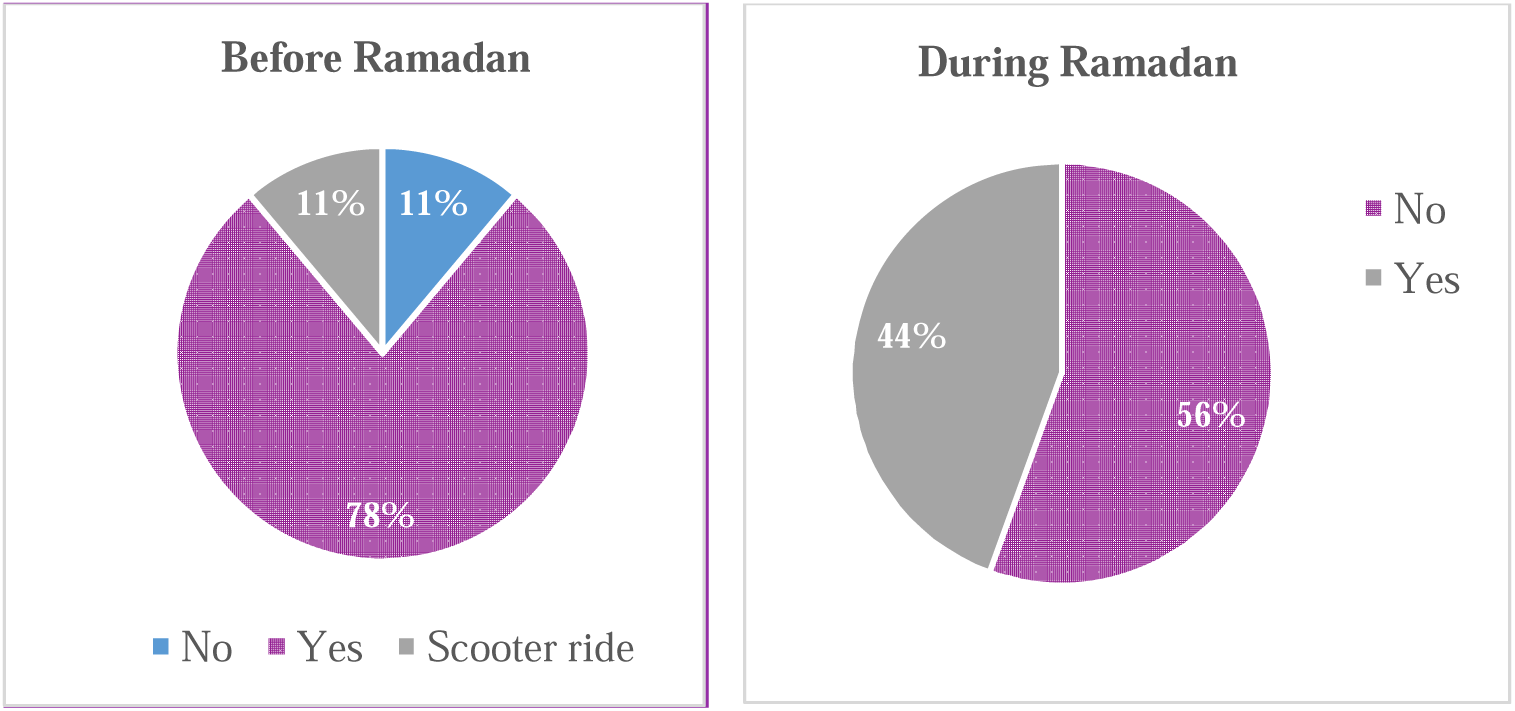
The percentages of the participants with T2D taking part in physical activities regularly before and during Ramadan (N=9). 44% (N=4); 56% (N=5); 78% (N=7); 11% (N=1).

**Figure 6:**
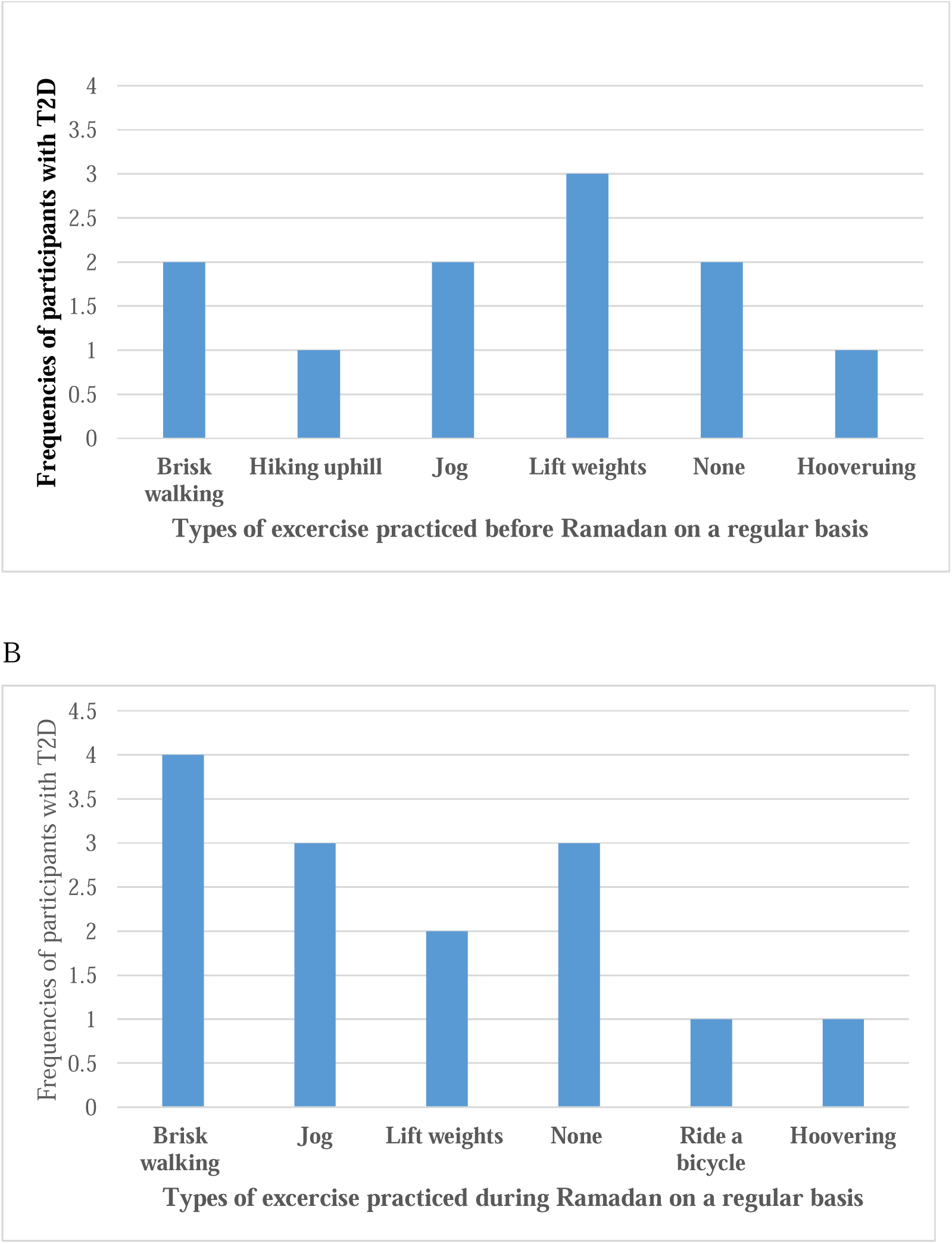
Different types of exercise practised by participants with T2D before Ramadan (A) and during Ramadan (B).

**Table 4** shows that the frequencies of the light exercise decreased among one third of the participants (N=3), from once a day before Ramadan to 2-4 times a week during Ramadan. Similarly, practising moderate exercise decreased from 2-4 times per week before fasting to never and 1-3 times a month during Ramadan. However, the frequency of vigorous exercise remained about the same before and during Ramadan, once a week at N=3. Furthermore, it was observed that 8 (89%) respondents reported that they were less active during the month of Ramadan, while about 6 (67%) respondents reported that they were more active before Ramadan (**Figure 7**).

**Figure 7:**
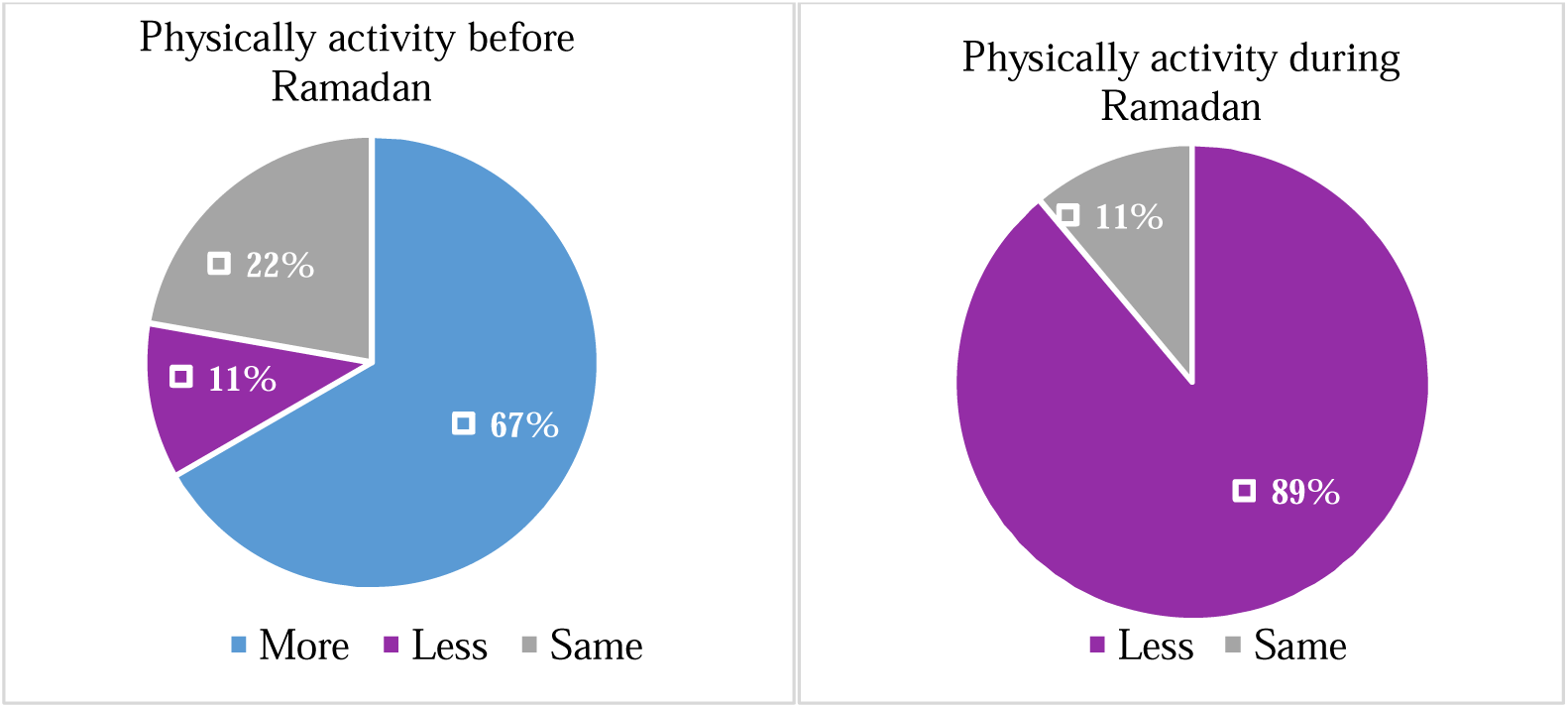
Comparison between the physical activities before and during Ramadan 2021 based on the respondents’ assessments (N=9). 67% (N=6); 22% (N=2); 11% (N=1); 89% (N=8).

**Table 4:** Comparison of the exercise patterns before and during Ramadan.

|  |  | Before Ramadan | During Ramadan |
| --- | --- | --- | --- |
|  |  | N | N |
| **Light exercise** | Never | 1 | 1 |
|  | 1-3/month | 0 | 2 |
|  | Once a week | 0 | 2 |
|  | 2-4/week | 4 | 3 |
|  | 5-6/week | 0 | 1 |
|  | Once a day | 3 | 0 |
|  | 2-3/day | 1 | 0 |
| **Moderate exercise** | Never | 2 | 3 |
|  | 1-3/month | 0 | 2 |
|  | Once a week | 2 | 1 |
|  | 2-4/week | 5 | 0 |
|  | 5-6/week | 0 | 2 |
|  | 2-3/day | 0 | 1 |
| **Vigorous exercise** | Never | 4 | 5 |
|  | 1-3/month | 1 | 0 |
|  | Once a week | 3 | 3 |
|  | 2-4/week | 1 | 1 |
| **Physically assessment** | More | 6 | 0 |
|  | Less | 1 | 8 |
|  | Same | 2 | 1 |

**Table 5** illus trate s the slee ping pattern before and during the month of Ramadan in T2D participants; 4 respondents reported that they fell asleep in <15 minutes during Ramadan, compared to 3 respondents before the month of Ramadan. Moreover, out of the 9 participants, 3 reported that they usually fall asleep in > 60 minutes before Ramadan, compared to only 2 respondents during the month of Ramadan. It was observed that participants went to bed for sleep at 4 am (median; range from 23:00–1:00) and got up at 9 am (median; range from 6:00–15:00). However, data indicated that sleeping hours at night and the total sleeping hours (day and night) were quite similar before and during Ramadan (**Table 5**). Respondents (N=3) reported that they usually have >8 hours of total sleep before Ramadan. This was reported by just one participant during the month of Ramadan. In addition, about half of the respondents (N=4, 45%) reported that the sleep quality was fairly bad, and one participant reported it as very bad during the month of Ramadan. On the other hand, before Ramadan, 3 respondents reported that the sleep quality was very good, and the other 6 respondents reported it as fairly good (N=2), fairly bad (N=2), and very bad (N=2) **(Figure 8**).

**Figure 8:**
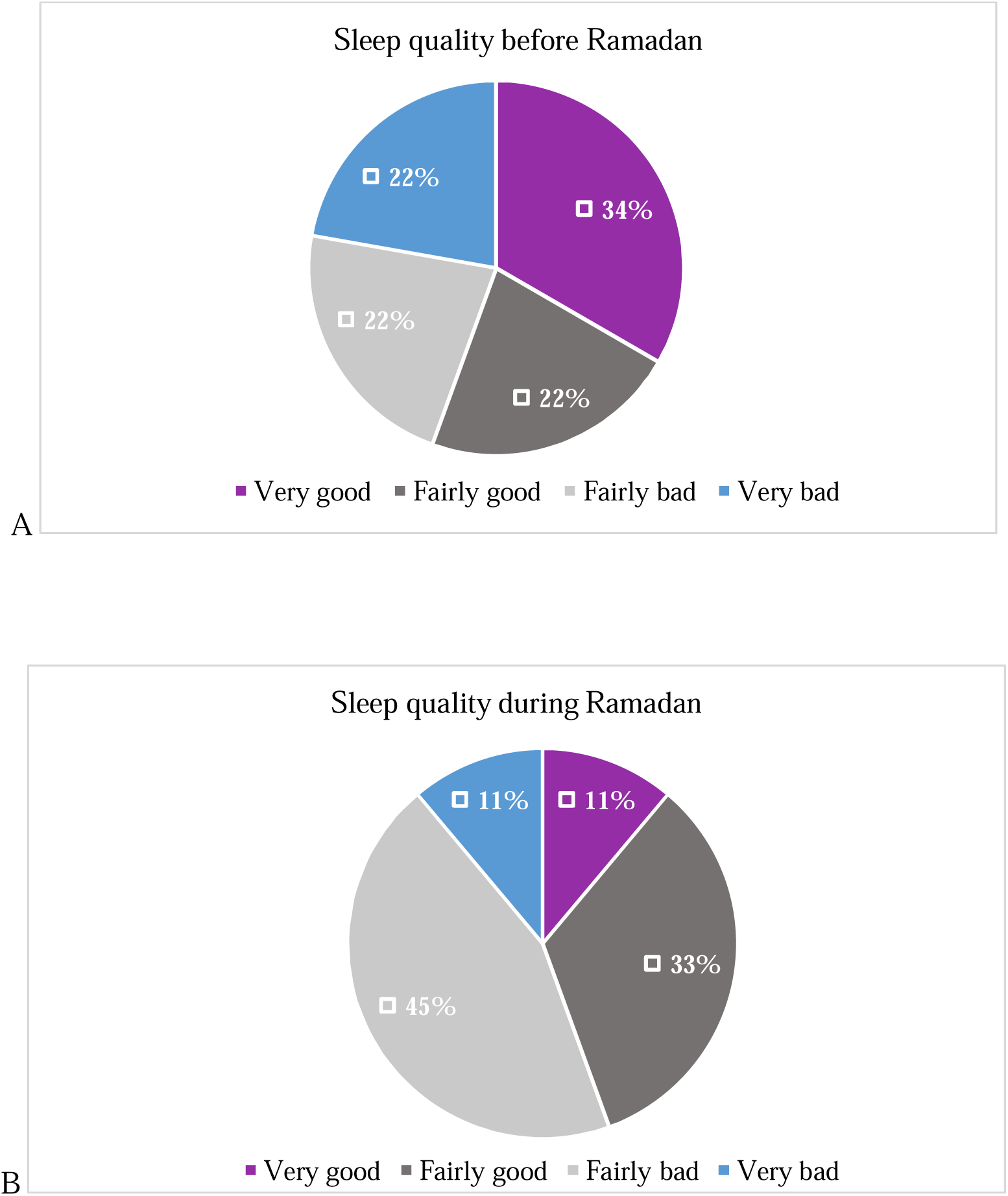
Comparison between the sleep quality before (A) and during (B) Ramadan among participants with T2D. 45% (N=4); 33% (N=3); 22% (N=2); 11% (N=1).

**Table 5:** Comparison between sleeping patterns before and during the month of Ramadan (N=9)

|  |  | Before Ramadan | During Ramadan |
| --- | --- | --- | --- |
|  |  | N | N |
| **Time to fall asleep each night** | < 15 minutes | 3 | 4 |
|  | 15-30 minutes | 2 | 2 |
|  | 31-60 minutes | 1 | 1 |
|  | >60 minutes | 3 | 2 |
| **Sleeping hours at night** | Less than 5 hours | 2 | 2 |
|  | 5-6 hours | 1 | 3 |
|  | 7-8 hours | 3 | 3 |
|  | > 8 hours | 2 | 1 |
|  | Depends on the night | 1 | 0 |
| **Total sleeping hours** | Less than 5 hours | 1 | 2 |
|  | 5-6 hours | 2 | 3 |
|  | 7-8 hours | 3 | 3 |
|  | More than 8 hours | 3 | 1 |
| **Sleep quality** | Very good | 3 | 1 |
|  | Fairly good | 2 | 3 |
|  | Fairly bad | 2 | 4 |
|  | Very bad | 2 | 1 |
| **Medicine to help with sleep** | No | 9 | 9 |
|  | Yes | 0 | 0 |

### Clinical characteristics for the participants before and after Ramadan 2021

**Table 6** shows that the mean body weight did not change after the month of Ramadan 2021 (p=0.343). Similarly, all the other parameters, including BMI, systolic and diastolic blood pressure, did not change significantly after the month of Ramadan. The level of HbA1c was slightly increased by -0.87 after Ramadan, but this was statistically not significant.

**Table 6:** Comparing the clinical and the anthropometric measurements before and after the month of Ramadan 2021

| | Mean ± SD | | P value |
| --- | --- | --- | --- |
|  | Before Ramadan | After Ramadan |  |
|  | Median (range) | Median (range) |  |
| Weight (Kg) (N=8) | 89.25 (68 - 122.5) | 89.23 (68-122) | 0.343 |
| BMI (kg/m<sup>2</sup>) (N=8) | 31.29 (25.59-45) | 31.47 (26.56-46.78) | 0.249 |
| SBP (mmHg) (N=4) | 123.50 (120-144) | 129.50 (119-135) | 1.000 |
| DBP (mmHg) (N=4) | 74.00 (69-84) | 76.50 (70-78) | 0.465 |
| HbA1c (N=4) | 57.63 (35-103) | 58.50 (36-89) | 0.715 |

P-value based on Related-Samples Wilcoxon Signed Rank Test; p-value significant at the level of 0.05. Data were collected about 2 months before Ramadan and 3 months after the month of Ramadan. BMI, body mass index; SBP, systolic blood pressure; DBP, diastolic blood pressure; HbA1c; Haemoglobin A 1c

### Impacts of the COVID-19 pandemic on the lifestyle of the participants before and during the month of Ramadan

Out of the 9 participants, just one respondent reported that he had a COVID-19 infection and a household with a COVID-19 infection before Ramadan, and he was willing to fast the month of Ramadan. Moreover, respondents reported that the COVID-19 pandemic was associated with weight gain before Ramadan (N=4, 44%); however, this was not reported during the month of Ramadan (**Figure 9**). Moreover, the vast majority (N=6, 67%) did not notice any change in body weight during the month of Ramadan, and one third reported (N=3, 33%) that the COVID-19 pandemic was associated with weight loss before and during Ramadan. A respondent who had a COVID-19 infection before Ramadan reported weight gain. Compared to the previous months of Ramadan, 5 (56%) participants reported that fasting during Ramadan 2021 was not stressful. However, 3 (33%) respondents reported that it was more stressful **(Figure 10**).

**Figure 9:**
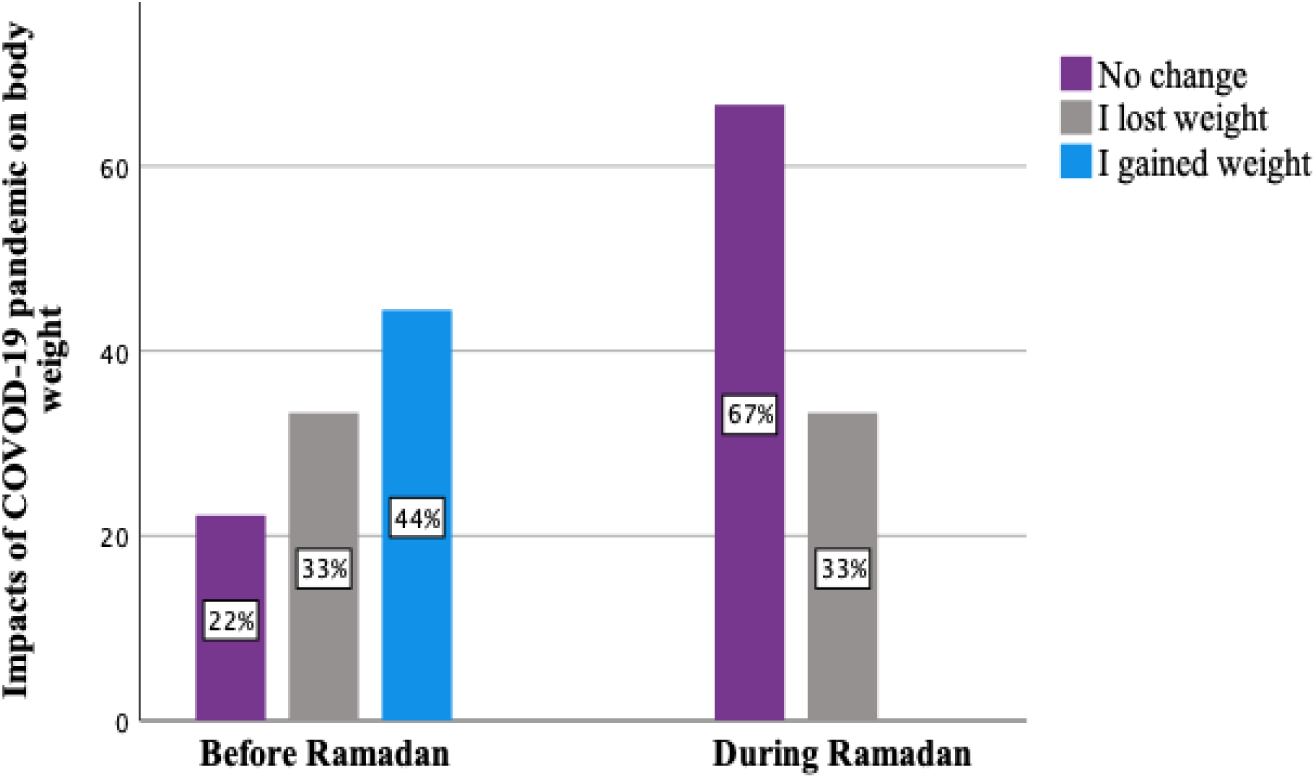
COVID-19 impacts on body weight before and during the month of Ramadan based on the participant’s evaluation. 67% (N=6); 44% (N=4); 33% (N=3); 22% (N=2).

**Figure 10:**
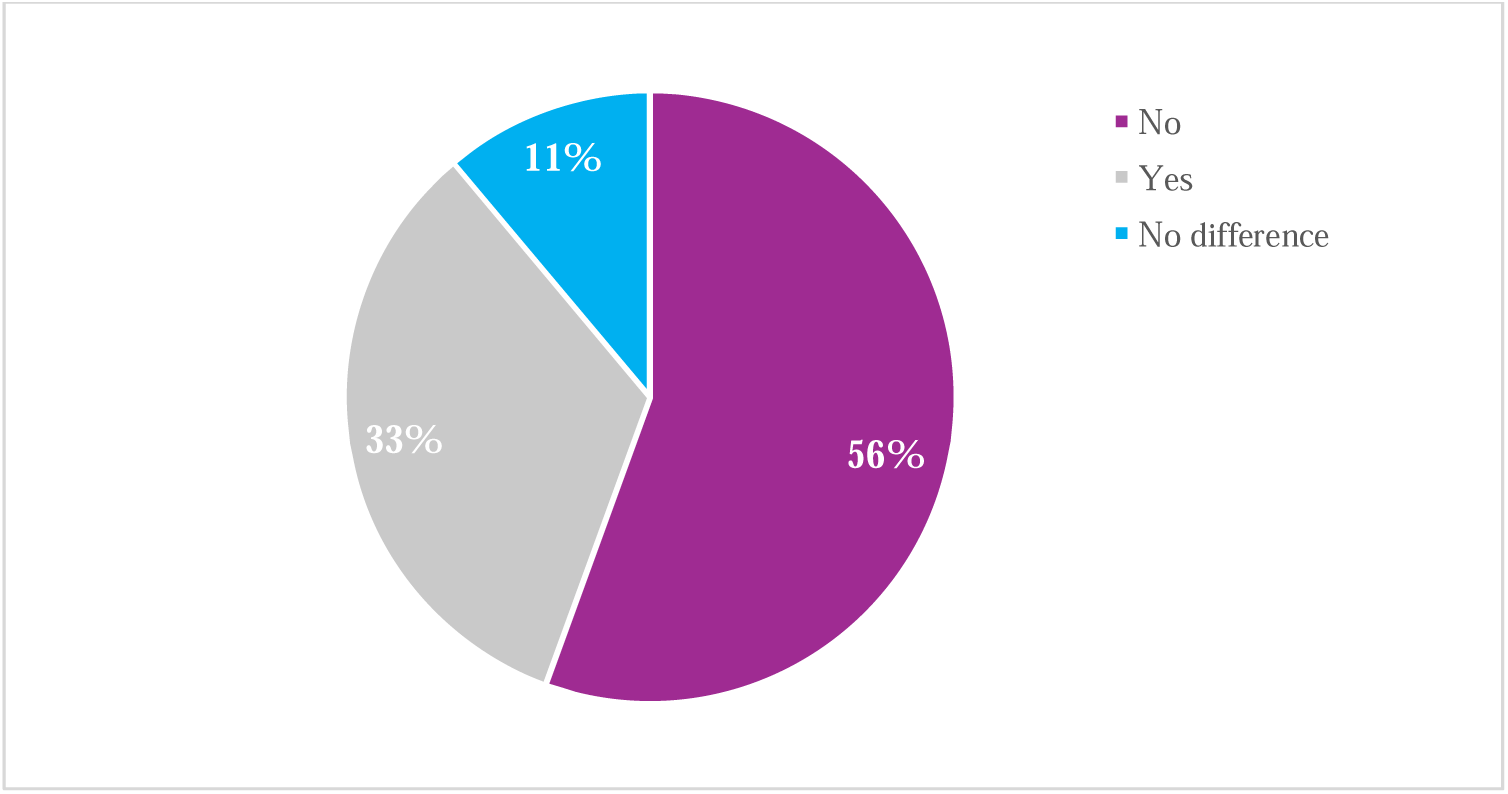
Participants’ assessment of the impact of the COVID-19 pandemic on the stress level during Ramadan 2021 compared to the month of Ramadan in previous years (pre-pandemic period) (N=9). 56% (N=5); 33% (N=3); 11% (N=1).

In addition, participants reported that the COVID-19 pandemic was associated with depression (N=2), emotional issues (N=3), family problems (N=2), and loneliness (N=2) before Ramadan. The reported frequencies of these issues slightly decreased during the month of Ramadan. Besides, participants (N=4) reported that there were no associated mental health issues during Ramadan compared to 3 participants before Ramadan (**Figure 11**). On the other hand, the frequencies of participants who reported that the COVID-19 pandemic was associated with stress were slightly decreased during the month of Ramadan compared to before Ramadan, at N=3 and N=4, respectively (**Figure 11**). One respondent reported anxiety before Ramadan, but this was not reported during Ramadan. However, financial problems (N=1) were reported only during Ramadan. Participants who had a COVID-19 infection before Ramadan experienced depression, stress, emotional issues, loneliness, and anxiety, and this was not reported during Ramadan (supplementary material).

**Figure 11:**
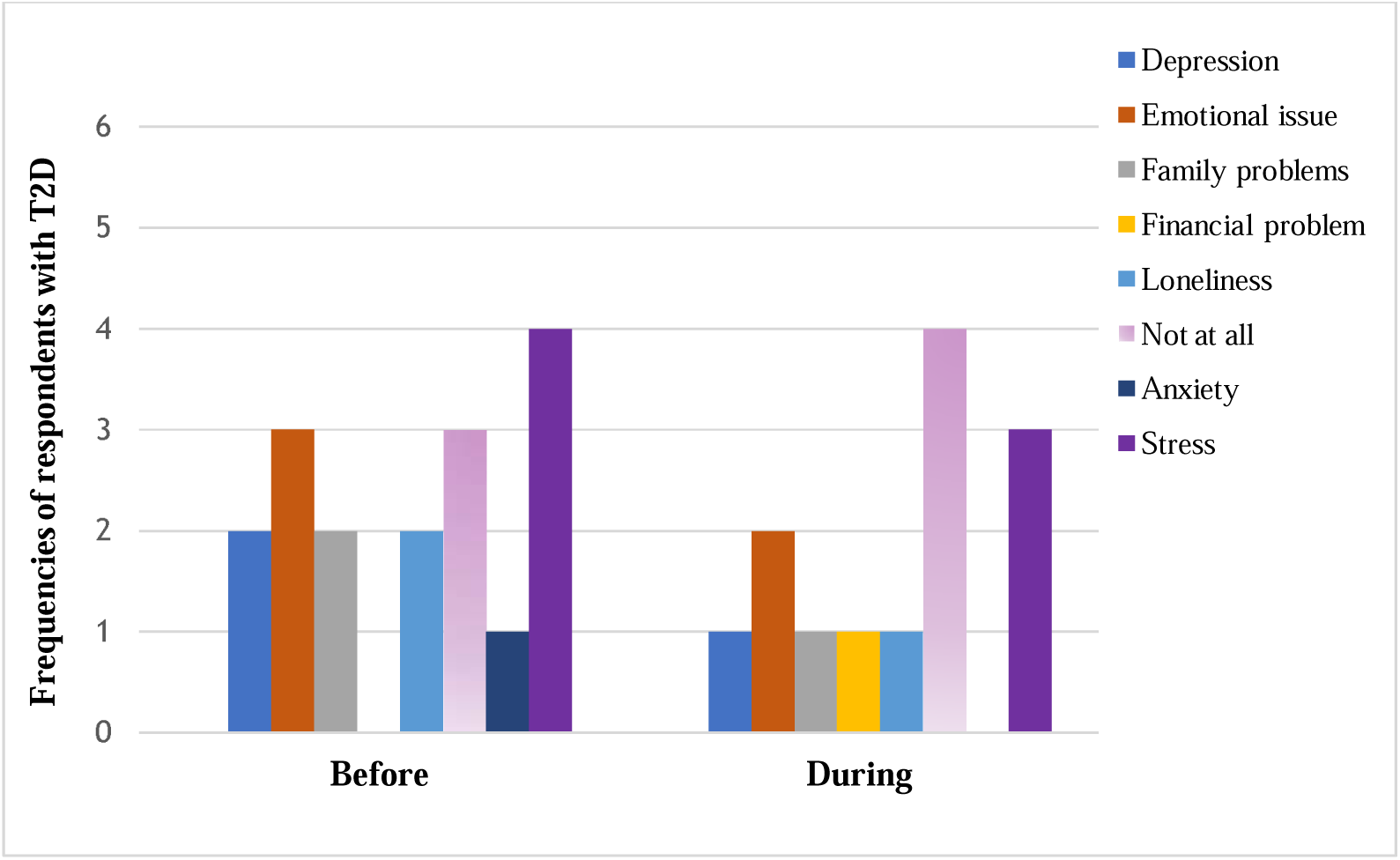
COVID-19-associated mental health issues among the respondents with T2D before and during Ramadan 2021.

**Figure 12** illustrates the relationship between the impacts of COVID-19 pandemic-related mental health issues on physical activities and body weight. Participants who had depression, stress, emotional issues, loneliness, and anxiety reported that they were more active before Ramadan. Moreover, respondents reported that depression, emotional issues, loneliness, and stress were associated with gaining weight before the month of Ramadan, and this association was not reported during Ramadan.

**Figure 12:**
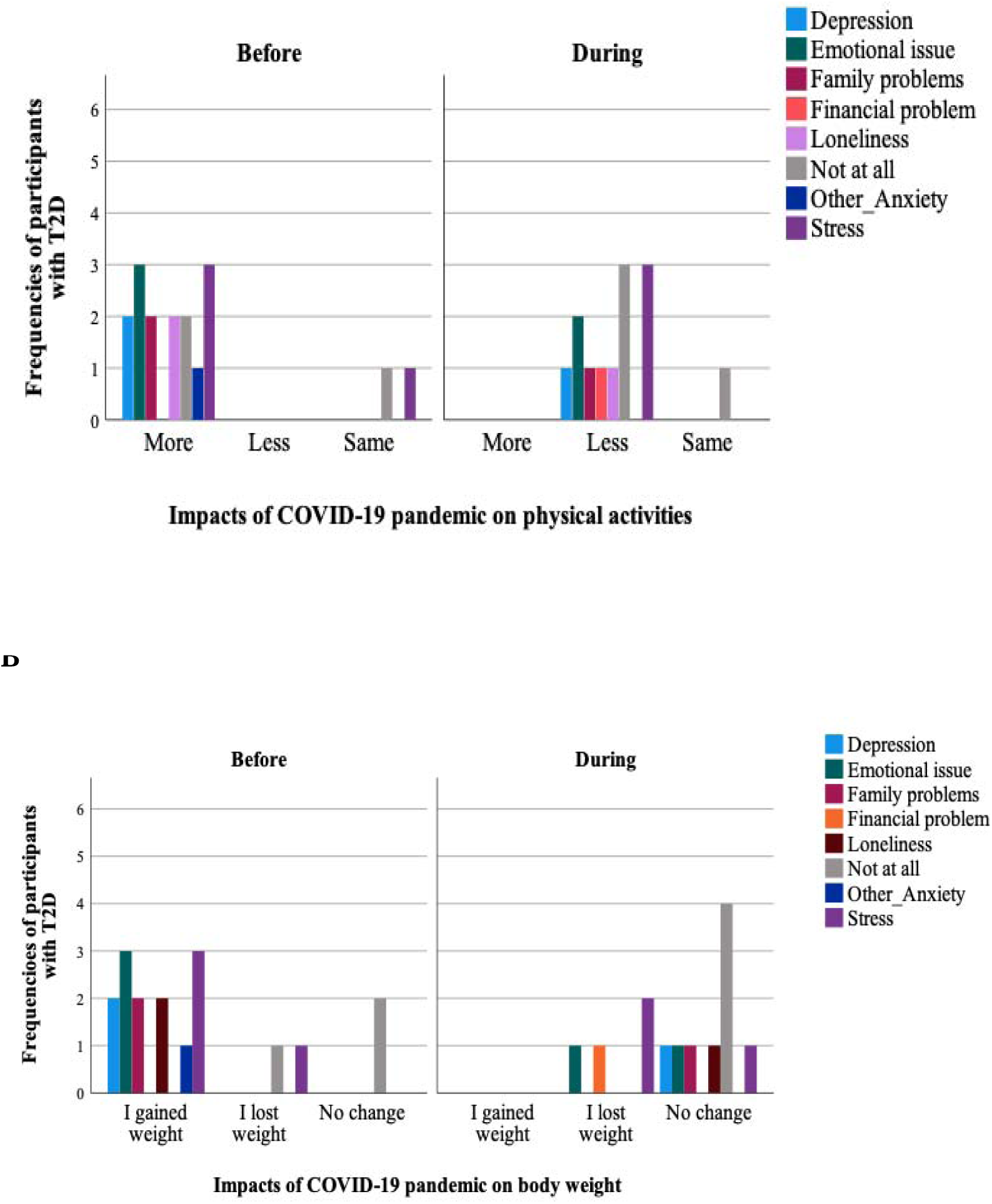
Associations between the impact of COVID-19-associated mental health problems on physical activity assessment (A) and body weight (B) among young T2D participants (N=9).

Participants (N=4) reported that the COVID-19 pandemic had restricted their access to medical services before Ramadan much less than before the pandemic, and the other 2 respondents reported that it was a little less than before. However, during Ramadan, more than half (N=5) reported that the general health care services were the same as before the pandemic period (**Figure 13**).

**Figure 13:**
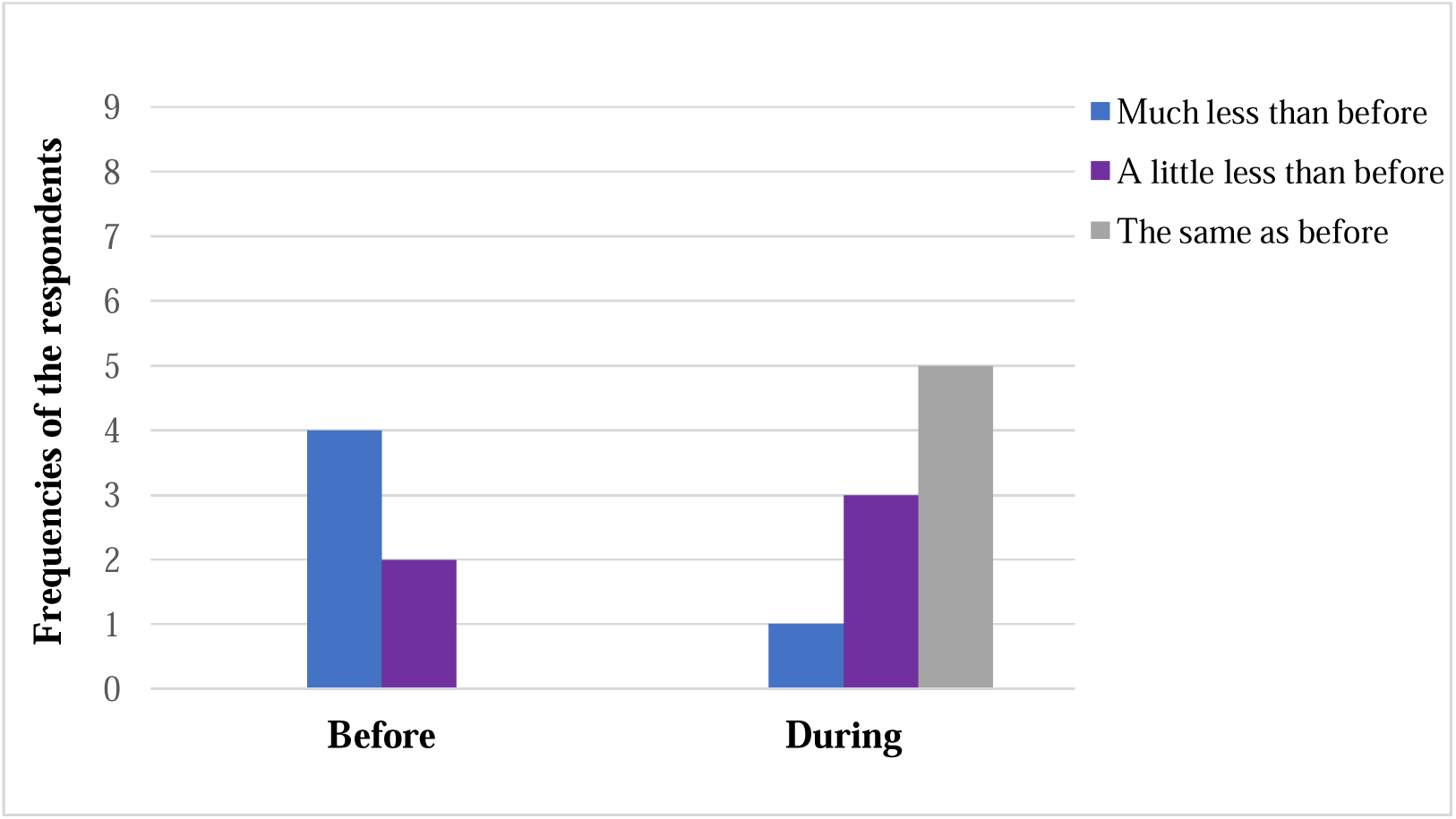
Impacts of the COVID-19 pandemic on access to general care before and during Ramadan 2021 (N=9).

Co mpa red to the mon th of Ra mad an in previous years (pre-pandemic period), 8 participants reported that they were less active during Ramadan 2021 (**Table 7**). Moreover, out of 9 respondents, 7 reported that the COVID-19 pandemic was not associated with longer sleep during Ramadan 2021. In addition, respondents (N=4) reported that fasting during the pandemic period was not associated with any complications compared to the previous months of Ramadan. Besides, participants (N=4) reported that their blood glucose slightly improved during RF, and one respondent reported that his diabetes greatly improved during fasting. No participant reported that their diabetes got worse during fasting, compared to RF in previous years during the pre-pandemic period (**Table 7**). Furthermore, respondents (N=5) reported that the COVID-19 pandemic had no impact on the quality of the foods consumed during Ramadan. Also, participants (N=6) reported that there was no problem with the availability of food during Ramadan during the pandemic period.

**Table 7:**
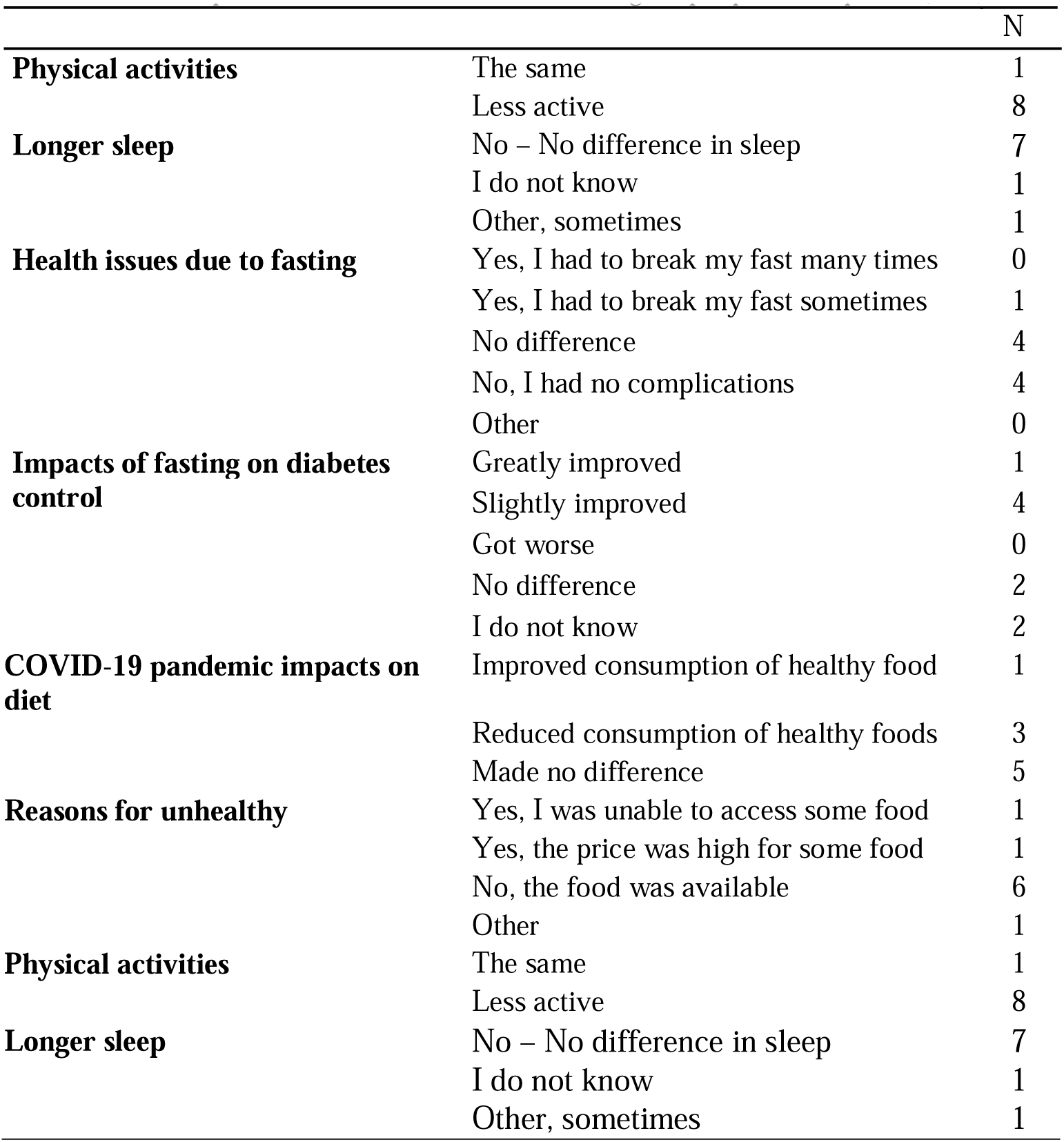
Impact of the COVID-19 pandemic on health issues and lifestyle sectors during Ramadan 2021 compared to the months of Ramadan during the pre-pandemic period (N=9).

Studying the relationship between variables using descriptive exploration and the Fisher exact test revealed that there was no relationship between physical activities and weight gain and/or weight loss before and during Ramadan in the pandemic period (**Figure 14**). Respondents (N=5) reported that they were less active and did not notice any change in body weight during Ramadan. Three respondents reported that they were more active and gained weight before Ramadan, but this was not reported during the month of Ramadan. However, the other 3 respondents who described themselves as less active during Ramadan reported weight loss.

**Figure 14:**
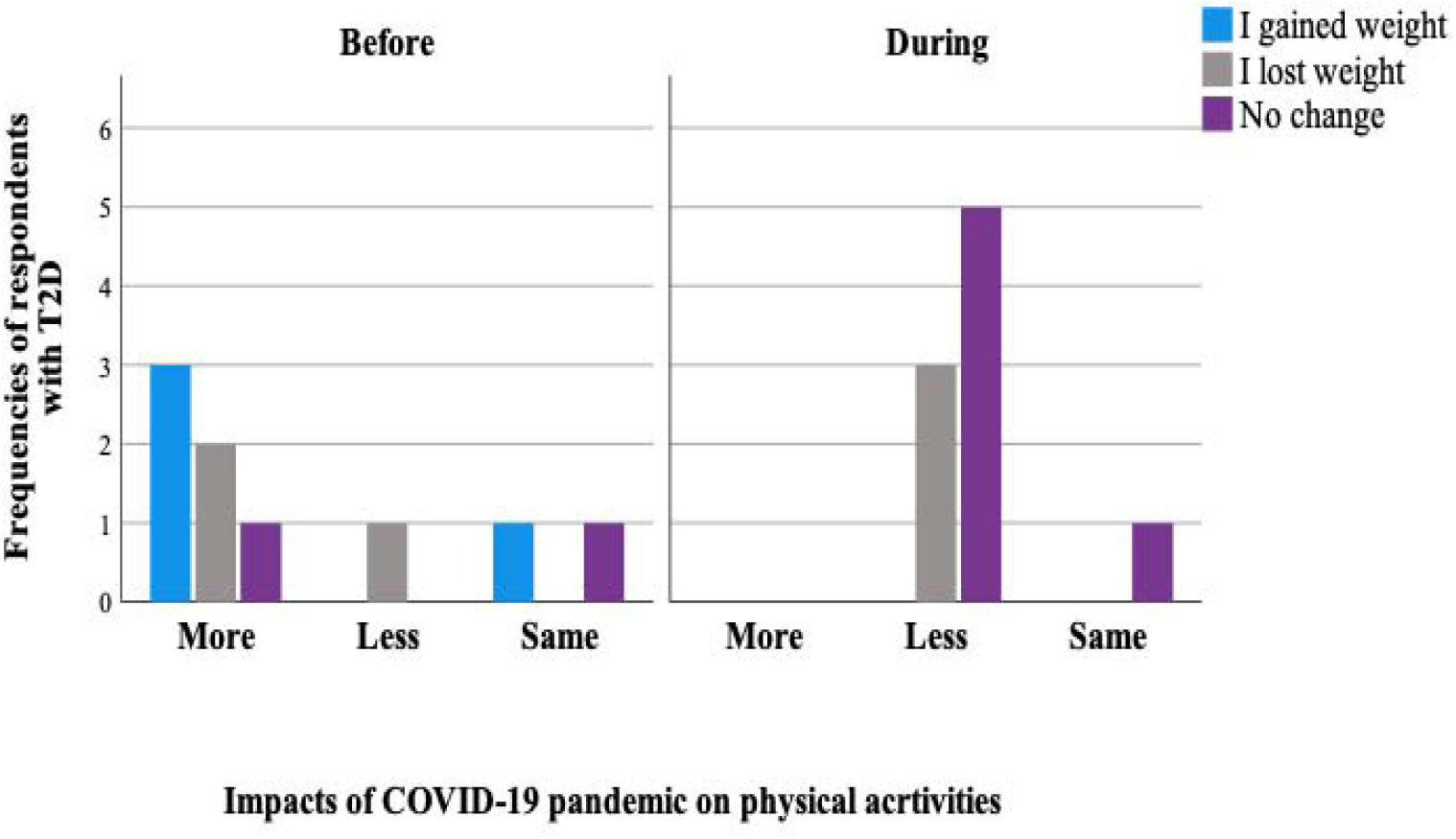
Association between the impact of the COVID-19 pandemic on physical activities and body weight (N=9).

## Discussion

This study provides evidence that children and young people with T2D in the UK fasted the month of Ramadan during the pandemic and did not experience any serious complications during fasting. The participants reported that they used to observe the month of Ramadan over the last few years because of the health benefits and feeling better during fasting. However, a few participants reported that social pressure had an impact on their decision to fast or not. Half of the study participants fasted the whole month of Ramadan. There was no change in their HbA1c, BMI, body weight, or blood pressure before and after Ramadan 2021. Similarly, the feasibility of fasting was reported among children/adolescents with T1D, even though the disease pathogenesis varied between the different types of diabetes [26]. It has been shown that there was no significant change in the level of biochemical parameters during fasting compared to before and after RF [26]. In addition, with appropriate training for parents, children/adolescents (> 8 years) with T1D can fast safely during the month of Ramadan [26, 27]. However, Alamoudi et al [28] reported that Saudi patients with T1D who fasted during the pandemic suffered from recurrent hypoglycaemia and hyperglycaemia, and the average fasting days were around 24 days. In addition, Zabeen et al [29] concluded that the COVID-19 pandemic was associated with minor impacts on young Bengali patients with diabetes (aged 14–24) with a non-significant rise in HbA1c after Ramadan.

It can be argued that fasting during Ramadan has a great influence on physiological/ biological mechanisms, and this could be related to the dietary patterns, including macro and micronutrients. Researchers showed that dietary patterns significantly changed during RF with increased protein and fat consumption in healthy young boys aged from 12 to 15 years old [30]. Moreover, they found that this contributed to the significant weight gain in the last week of Ramadan and two weeks after Ramadan. In addition, they conducted this study based on a 24-hour recall supported by digital records [30]. Furthermore, dietary patterns during Ramadan are highly affected by cultural habits. For example, a substantial rise in the consumption of carbohydrate-based foods and fat has been reported among healthy Algerian young adults [31]. Herein, similarly, data showed increased consumption of desserts with increased consumption of fruits and vegetables by young study participants with T2D; however, the weight gain was not reported.

Moreover, a prospective study conducted among healthy adolescents (mean age of 15 years, N=366) in Ghana reported that the dietary patterns were remarkably altered during the month of Ramadan [32]. They found a significant reduction in the consumption of fast food with increased intake of fruits, vegetables, and a variety of foods, and this was associated with transient weight loss during fasting. Moreover, they reported that about half of the participants practised fasting after the month of Ramadan [32]. Therefore, it could be argued that the presence of diabetes has little impact on dietary habits, and patients behave similar to healthy people. Assessment of diet intake and frequent evaluation of the amount of consumed nutrients are essential and part of diabetes management [33]. However, nutrition management among children/adolescents with diabetes is associated with many challenges and low compliance. The young participants with T2D investigated in this research reported a diet between unhealthy and average before Ramadan. Interestingly, during RF, one-fifth of healthy diet consumption emerged. This could explain the maintenance of the same weight during fasting among study participants who are between obese and overweight.

On the other hand, most of the participants in this study reported gaining weight during the period before fasting, and it was indicated that this was mainly related to the COVID-19 pandemic’s negative consequences. Poor adherence to the recommended diet and physical activities in people with obesity during the pandemic has been reported [34]. Moreover, it could highlight that the quarantine consequences, including stress, anxiety, and restriction of physical activities, were associated with weight gain and deterioration in sleeping and diet patterns in adult patients with diabetes (T1D and T2D) [35, 36]. Several studies reported that physical activity decreased during the months of Ramadan among adult patients with T1D and T2D [37, 38]. Consistently, this was observed in the current research study among the young people with T2D. Interestingly, another study among adults with T2D found that RF has no impact on physical activities, and it was observed that the level of activity did not vary from the period before Ramadan [39]. This could be related to the natural variations in the study participants included in different studies. Furthermore, a recent study conducted by Abdelmalek et al [40] reported that exercising during RF was associated with significant weight loss among healthy adolescents with obesity [40]. It has been suggested that education about the maintaining physical activities and healthy diet during fasting is paramount [41].

Investigating the impacts of RF on sleeping patterns among Saudis with T2D and T1D (>20 years old), it was found that the total number of sleeping hours has not changed during the months of Ramadan [42]. Study participants reported that they had their sleep time mainly during the daytime instead. Moreover, a reduction in sleep duration and quality has been reported among young, healthy athletes during Ramadan [43]. Consequently, this could harm mental health by increasing stress and anxiety. However, it has been shown that the severity of depression was reduced during the month of Ramadan in adult patients with diabetes [44]. Likewise, this was seen and reported by the young T2D participants studied in this research. During the COVID-19 pandemic, children with T1D were found to be noticeably impacted by the quarantine measures compared to adolescent patients [45]. It was found that adolescents showed the ability to accommodate, and they were engaged in several physical activities during the pandemic period. Importantly, evidence indicates that the incidence of depression among patients with T2D is remarkably high and is related to uncontrolled blood glucose and diabetes comorbidities [46, 47]. Moreover, this has been observed among the children and young adults with T1D as well [48]. Thus, improvement in depression levels during fasting could have a great impact on the quality of life of individuals with diabetes and the disease prognosis.

Therefore, close communication with medical professionals before Ramadan could have a great impact on maintaining blood glucose control during fasting [49]. This could be achieved by running pre-Ramadan clinics for all patients who are willing to fast during Ramadan, in terms of adjusting diet, emphasising the maintenance of physical activities, and improving sleep quality during fasting [50]. However, this is not common practice in Western countries, for instance, in the UK. Recently, the official website of Diabetes UK has been providing advice and guidance on the importance of following the best-agreed plan with a medical professional before fasting [51]. This is usually shared with patients as an information leaflet in some centres. Yet, it could be mentioned that this approach may not be suitable for young people with diabetes, and direct face-to-face or virtual pre-Ramadan education sessions with the patients and their parents are necessary and more effective.

## Limitations of the study

This study has several limitations, including a small sample size of the studied participants. As a result, some statistical analyses were not possible to apply to identify the differences between some categorical and continuous variables. In addition, the relationship between variables could not possibly be assessed statistically. Furthermore, it was not possible to introduce a face-to-face questionnaire as it was conducted during the COVID-19 pandemic period. Consequently, this could have an impact on the quality of diet assessment as it was based on the recorded food frequencies and not on a specific food portion.

## Conclusion

The findings of this research have contributed to current knowledge by providing data on young patients with T2D who chose to perform RF during the COVID-19 pandemic in 2021. These study participants fasted safely during Ramadan. Fasting during the COVID-19 pandemic circumstances was not stressful for most participants and improved patients’ mental health with no significant change in body weight or blood glucose parameters after the month of Ramadan. Dietary patterns were variable, and the sleeping pattern was fairly bad during Ramadan; this was not related to the impact of the COVID-19 pandemic. Although there was a clear message from the healthcare professionals on the importance of keeping physically active during the pandemic, young patients with T2D were physically inactive and clearly impacted by the pandemic restriction measures. Soon, much larger studies will be necessary to determine the impacts of RF on young people with T2D. This could be carried out through prospective studies or retrospective data from patients in hospitals and medical centres, especially in terms of their adherence to medication, blood glucose control, and lifestyle patterns during fasting.

## Data Availability

All data produced in the present work are contained in the manuscript

## Acknowledgments

This work would not have been possible without the willingness of the study participants and their parents. Our deepest gratitude to all the staff who are working on the research and innovation administrators in the three NHS trusts, Leicester, Bradford, Birmingham, and De Montfort University. They are very much appreciated for their effort to mitigate the risk of infection and for providing the best support to conduct a great part of this research during the COVID-19 pandemic. All co-authors would like to sincere thanks the research coordinator, Rutger Clarke from Bradford Teaching Hospitals NHS Foundation Trust, and Dr. Douglas Gray, head of the Faculty of Health and Life Sciences Research Ethics Committee (FREC) at De Montfort University, for their great support and guidance in understanding the process of setting up the study and obtaining the research ethical approvals from the Research Ethics Committee (REC), the Health Research Authority (HRA) in the UK.

## Conflict of Interest

The authors declare that the research was conducted in the absence of any commercial or financial relationships that could be construed as a potential conflict of interest.

## Notes

### Competing Interest Statement

The authors have declared no competing interest.

### Funding Statement

Funding for this research study was provided by PhD scholarship awarded to the first author Dr Hala Elmajnoun.

### Author Declarations

Ethical approvals were obtained from the Health Research Authority (HRA) and from the Health and Life Science Faculty Research Ethics Committee at De Montfort University.

