## Supplementary material for "Effects of Ramadan Fasting on Children and Young Adults with Type 2 Diabetes during the COVID-19 Pandemic: A Prospective Study": Supplementary Material_RF and T2D prospective study .pdf

#### Participants information sheets (PISs)

PIS for 12-15 years old with T2D

PIS for parents

PIS for 18+ years with T2D

PIS for 16-18 years with T2D

#### Study questionnaires

A questionnaire for 12-15 years with T2D before Ramadan

A questionnaire for 12-15 years with T2D after Ramadan

A questionnaire for 16-24 years with T2D before Ramadan

A questionnaire for 16-24 years with T2D after Ramadan

#### Study protocol

Prospective study protocol

#### Research flyer

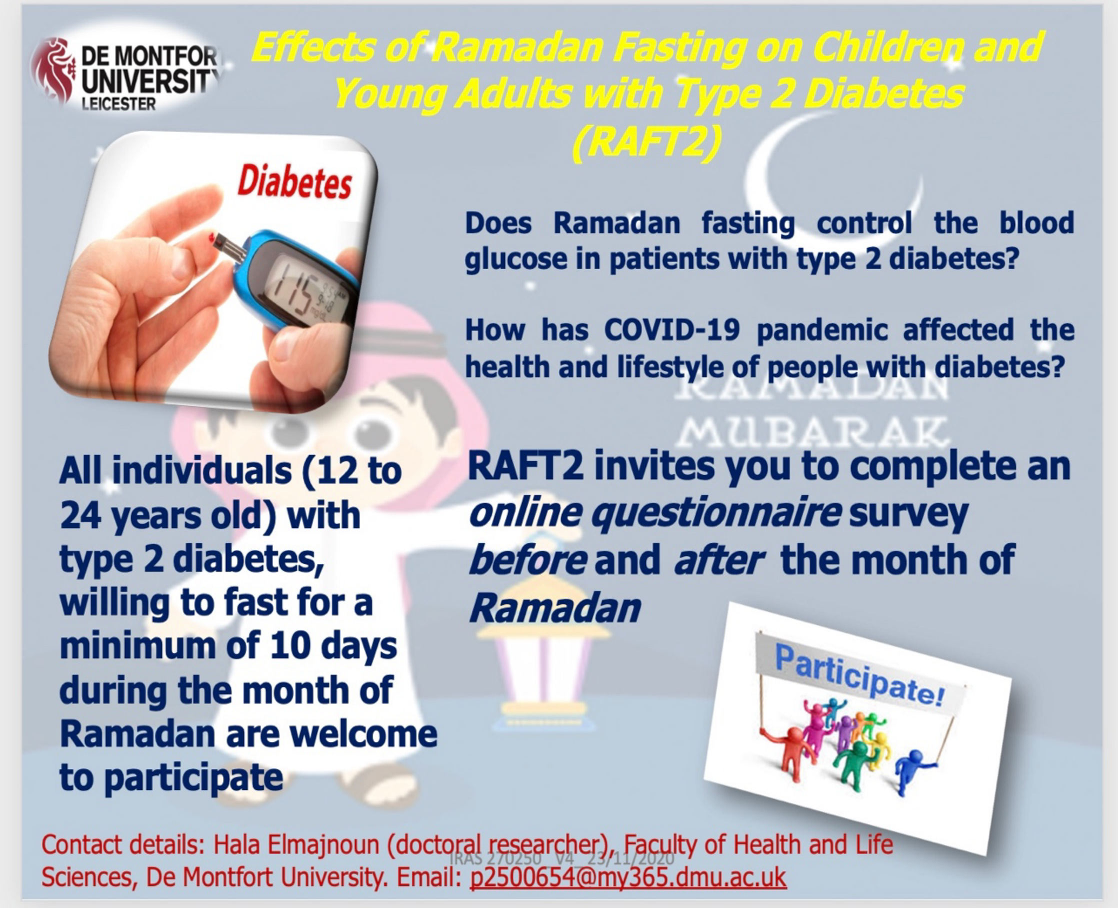

### Approval letter for the study

**Wednesday, February 24, 2021 at 11:58:21 Greenwich Mean Time**

**Subject:** IRAS PROJECT ID 270250, REC Reference 20/LO/0609 Confirmation of favourable opinion for substantial amendment
**Date:** Wednesday, 23 December 2020 at 10:00:32 Greenwich Mean Time
**From:**
**To:**, Hala Elmajnoun
**Attachments:** IRAS_270250_Favourable_opinion_of_a_substantial_amendment_23_12_2020.pdf

Dear Professor Haris

| | |
|---|---|
| **IRAS project ID:** | 270250 |
| **REC reference:** | 20/LO/0609 |
| **Short Study title:** | Ramadan fasting in patients with type 2 diabetes (RAFT2) |
| **Date complete amendment submission received:** | 10 December 2020 |
| **Amendment No./ Sponsor Ref:** | Ref 3386 |
| **Amendment Date:** | 23rd November 2020 |
| **Amendment Type:** | **Substantial** |
| **Outcome of HRA Assessment** | **This email also constitutes HRA and HCRW Approval for the amendment, and you should not expect anything further.** |

I am pleased to confirm that this amendment has been reviewed by the Research Ethics Committee and has received a Favourable Opinion. Please find attached a copy of the Favourable Opinion letter.

**HRA and HCRW Approval Status**

As detailed above, **this email also constitutes HRA and HCRW Approval for the amendment**. No separate confirmation of HRA and HCRW Approval will be issued.

**User Feedback**

The Health Research Authority is continually striving to provide a high quality service to all applicants and sponsors. You are invited to give your view of the service you have received and the application procedure. If you wish to make your views known please use the feedback form available on the HRA website: http://www.hra.nhs.uk/about-the-hra/governance/quality-assurance/.

If you require further information, please contact me.

Kind regards

**Laura Howe**
**Approvals Officer**
Skipton House | 80 London Road | Health Research Agency | SE1 6LH
**T.** 02071048387
**E.**
**W.** www.hra.nhs.uk

Sign up to receive our newsletter HRA Latest.
